# The Persistence of Vaccine Hesitancy: COVID-19 Vaccination Intention

**DOI:** 10.1101/2020.12.16.20248139

**Authors:** Jagadish Thaker

## Abstract

Building public trust and willingness to vaccinate against COVID-19 is as important as developing an effective vaccine. However, a significant minority of the public are unwilling or hesitant to take a COVID-19 vaccine, when available. A nationally representative sample survey (*N*=1040) was conducted in July 2020 in New Zealand to identify factors associated with COVID-19 vaccine intention. Trust in experts and general vaccine hesitancy were significantly associated with COVID-19 vaccine intention. A communication campaign from trusted scientific experts, with information that addresses prevailing concerns about vaccines, is likely to help increase COVID-19 vaccine uptake.

---

The coronavirus disease 2019 (COVID-19) pandemic is continuing to have an unprecedented impact on the public health and the global economy. Over 35.5 million cases and 1 million deaths have been reported in 227 countries and territories. New Zealand has been one of the few countries that is largely successful in containing the COVID-19 spread so far, due to tough border controls and contact tracing (World Health Organization, 2020).

An unprecedented scientific endeavor is underway to develop a vaccine against the severe acute respiratory syndrome coronavirus-2 (SARS-CoV-2), the virus that causes COVID-19. Just as important as developing a COVID-19 vaccine is to develop public enthusiasm for vaccination. Public attitudes towards vaccination can shift and it is important to gauge public perceptions of COVID-19 vaccine for high vaccination uptake when a COVID-19 vaccine becomes available (de Figueiredo et al., 2020; Fadda et al., 2020; Palamenghi et al., 2020; Verger & Dubé, 2020).

This study applies the theory of planed behavior (Ajzen, 1991), which states that attitudes play a key role in shaping behavioral intentions, which in turn, serve as a proximal predictor of behaviors. Other independent predictors of behavioral intention include subjective norms and behavioral control. Attitudes are defined as positive or negative assessment towards an object or an issue (Binder et al., 2009; also see Fazio, 1986). Attitudes towards vaccines can be defined as expression of support or hesitancy across different vaccines (e.g., Yaqub et al., 2014). While a number of studies assess attitudes towards vaccines, they have several limitations, including use of few measures that are not validated (Krishna, 2018; Xiao, 2019; Lin et al., 2020), student samples and limited number of studies outside the US and European context (see Larson et al., 2016 for review). Even recent studies on COVID-19 vaccine intentions use only a few items to measure vaccine hesitancy (Cadeddu et al., 2020; Detoc et al., 2020).

Vaccine hesitancy is one of the 10 most important health threats in the world today (World Health Organization, 2019). Vaccine hesitancy can be defined as reluctance or refusal to vaccinate despite its accessibility (MacDonald, 2015). Public health campaigns must address vaccine hesitancy long before actual refusal takes place, thereby increasing public confidence and inoculating the public against misinformation about vaccines. While most studies on vaccine hesitancy focus on subpopulations such as parents and caregivers or specific vaccines (e.g., HPV), a generalized vaccine hesitancy scale will better help understand the public unmet need for health information (Larson et al., 2015; Luyten et al., 2019). Previous attempts to develop a generalized vaccine hesitancy scale have been limited with only one study clarifying the dimensions of vaccine hesitancy among general public (Luyten et al., 2019).

The current study examined factors associated with the intention to get a COVID-19 vaccine using data from a national sample survey of adults in New Zealand. Given the novelty of the disease, apart from demographic correlates, other factors tested in the study are vaccine hesitancy (Luyten et al., 2019; Shapiro et al., 2018), trust in scientists (Cadeddu et al., 2020; de Figueiredo et al., 2020; Justwan et al., 2019; Larson et al., 2018; Palamenghi et al., 2020), and knowledge about COVID-19 origin, symptoms, spread, and preventive behaviours (Reiter et al., 2020). Trust in scientific experts is central in several science communication and health communication studies (e.g., Hmielowski et al., 2013; Malka et al., 2009; Poortinga & Pidgeon, 2003; Siegrist et al., 2005; Slovic, 1993) as information from trusted sources reduce cognitive effort and provides mental short-cuts to help individuals make decisions, particularly about highly uncertain issues, such as a new vaccine. Empirical evidence on the role of knowledge in shaping public acceptance of vaccines is mixed (Motta et al., 2018), therefore essential to study in the context of potential new vaccines. Findings of this study can help guild theory and practice of public health campaigns for COVID-19 vaccination uptake.

## Method

### Study design

A nationally representative sample survey of the New Zealand adults was conducted between June 26 to July 13, 2020, after the country was briefly at the Alert Level 1, with fewer restrictions. The web-based survey was fielded by Qualtrics, an international survey agency. A stratified random sampling plan was followed to match the official Census estimates. The average time to complete the survey was 22 minutes. Ethics approval to conduct the survey was received from the Massey University Human Ethics Committee (Ethics Notification Number: 4000022852 by Professor Craig Johnson, Chair, Human Ethics Chairs’ Committee and Director, Research Ethics). Participants provided informed consent after reading brief aims of the survey about public opinion about current issues facing the country and the world.

### Measures

#### COVID-19 Vaccination Intention

Following recent surveys (O’Keefe, 2020; Reiter et al., 2020), respondents were asked, “If a coronavirus vaccine were available soon, what would you do? - I will get vaccinated against the coronavirus when one becomes available.” Response options included “yes” and “no.”

#### Knowledge about COVID-19

Eleven questions measured dichotomously (*True* or *False*), were used to compute knowledge about COVID-19. These questions started with a prompt, “To the best of your knowledge, which of the following statements true or false.” All scientifically accurate statements were coded 1 compared to 0. Eight statements were scientifically accurate relating to symptoms (dry cough (*True*, 1, 87%), fever (93%)), spread (“the coronavirus can be spread by people who do not show symptoms,” *Yes*, 1, 93%), protection (frequent hand washing, 97%), avoiding large gatherings (92%), 6-feet distance (74%), and cure (“there is currently no cure for the coronavirus, 81%). Five statements were scientifically inaccurate, including on impact on elderly (“only elderly people get infected,” *False*, coded as 1, 94%), and protection (“Hydroxychloroquine can prevent or kill coronavirus,” *False*, 1, 84%; “antibiotics can prevent or kill the coronavirus,” *False*, 1, 84%; “exposure to sun or extreme heat can prevent or kill the coronavirus,” *False*, 1, 76%). Similarly, the respondents who correctly identified conspiracy theories as false were coded as 1 (“5G towers are spreading coronavirus,” *False*, 1, 93%; “Bill Gates may have created the coronavirus to profit,” *False*, 1, 90%; “coronavirus was created in a lab,” *False*, 1, 66%). The scientifically accurate answers were summed to create an index of knowledge about COVID-19 (*M* = 9.58, *SD* = 1.56; KR-20 (Kuder-Richardson Formula 20) = .58).

#### Vaccine Hesitancy Scale (VHS)

Following previous studies that validated vaccine hesitancy scale (Larson et al., 2018; Luyten et al., 2019; Shapiro et al., 2018), attitudes towards vaccination was measured using 14 items using a 5-point scale, from *strongly disagree* to *strongly agree* with *neither agree nor disagree* serving as a mid-point. The questions followed a prompt, “Regarding vaccines, how much do you agree or disagree with following statements.” These items include previously validated 9-item Vaccine Hesitancy Scale (Luyten et al., 2019): one-item, “I do not need vaccines for diseases that are not common anymore,” was not used as two recent studies found the item to be unreliable (Luyten et al., 2019; Shapiro et al., 2018). In addition, six new items sourced from systematic reviews about vaccine hesitancy were tested: “Vaccines cause diseases” (Gidengil et al., 2019), “Government over hypes the need for vaccines,” (Gidengil et al., 2019), “Corporations manufacturing vaccines only care for profit,” (Gidengil et al., 2019), “I am uncomfortable getting a vaccine that was rushed into production” (Hoffman, 2020; Seale et al., 2010), and “I feel uncomfortable getting vaccinated” (Maisonneuve et al., 2018).

#### Trust in Scientific Experts

Trust in scientific exerts was measured using four items starting with a prompt, “How much do you trust or distrust the following organizations or people as a source of accurate information?” Using a 5-point response scale, with *strongly distrust* to *strongly trust*, respondents were asked how much they trust in (1) scientists (*M*= 3.99, *SD*=1.09), (2) medical experts (*M*= 4.14, *SD*=1.02), (3) infectious disease experts (*M*= 4.11, *SD*=1.02), and (4) your doctor or GP (general practitioner) (*M*= 4.16, *SD*=.94). The four items were added to compute an index of trust in scientific experts (*r*’s ranged from .47 to .75, *p*<.001; *α* = .87, Kaiser-Meyer-Olkin measure = .82, Bartlett’s test of sphericity (*χ*2 (6) = 2151.39, *p* < .001)).

#### Demographic Characteristics

The survey assessed a range of demographic variables (see Table 1).

**Table 1.** Demographic Characteristics of the Sample

|  | *N* (unweighted) | % (unweighted) | % (weighted) | % Census Estimate |
| --- | --- | --- | --- | --- |
| Total | 1040 | 100 | 100 | 100 |
| *Sex* |  |  |  |  |
| Female | 609 | 58.6 | 51 | 50.6 |
| Male | 431 | 41.4 | 49 | 49.3 |
| *Age* |  |  |  |  |
| 18-25 | 189 | 18.2 | 14 | 14 |
| 26-35 | 220 | 21.2 | 18 | 18 |
| 36-45 | 175 | 16.8 | 16 | 16 |
| 46-55 | 163 | 15.7 | 18 | 18 |
| 56-65 | 127 | 12.2 | 15 | 15 |
| 66 and above | 166 | 16 | 19 | 19 |
| *Education* |  |  |  |  |
| No qualification | 96 | 9.2 | 19 | 18.19 |
| Level 1 to Level 6 diploma | 577 | 55.5 | 54 | 51.10 |
| Bachelor's degree or higher | 367 | 35.3 | 27 | 24.82* |
| *Ethnicity* |  |  |  |  |
| European New Zealander | 648 | 62.3 | 61.5 | 64 |
| Māori | 139 | 13.4 | 16.3 | 17 |
| Pasifika | 50 | 4.8 | 7.7 | 8 |
| Asian or other | 203 | 19.5 | 14.4 | 15.1 |

*Note:* *N*=1040. Data was weighted on sex, age, ethnicity, and education. The census estimates
according to 2018 census (https://www.stats.govt.nz/2018-census/). \* Percentages do not add to
100% as some responses were unidentifiable or not stated in the Census.

### Data analysis

The data was weighted, post-survey, on gender, age, education, and ethnicity to match the New Zealand census estimates. Weights ranged from 0.47 to 3.30, with a mean of 1.21, median of 0.98, and a standard deviation of 0.63. 95% of the weights fall between 0.58 and 2.15. To assess the dimensions of the 14-item vaccine hesitancy scale, the dataset was randomly and evenly split to test and validate the scale. Relative risk regression models with robust standard errors (Zou, 2004) were used to identify correlates of COVID-19 vaccine intension, as they are more intuitive than odds ratios and is a common method in public health research. Data were analyzed using IBM SPSS version 25 (IBM Corp., Armonk, NY).

## Results

### Participant Characteristics

Two-thirds (74%) of the respondents said they intend to get vaccinated against COVID-19 when one becomes available and a quarter (26%) said they do not intend to do so.

### Vaccine Hesitancy Scale

The sample was randomly split into half to test the dimensions of vaccine hesitancy scale. Exploratory factor analysis for the 14-items with the first half of the sample indicated a two factor solution, similar to prior studies (Luyten et al., 2019; Shapiro et al., 2018) (see Table 2). Kaiser-Meyer-Olkin measure of sampling adequacy was .93, above the commonly recommended value of .6, and Bartlett’s test of sphericity was significant (*χ*2 (91) = 5224.38, p < .001). These two factors were labelled as “lack of confidence” factor, accounting for 49.65% of the variance, and “risks” factor, accounting for 17.49% variance, based on previous studies (Luyten et al., 2019; Shapiro et al., 2018). These two factors accounted for 67.15% of the common variance of the scale.

**Table 2.** Factor Analysis of the Vaccine Hesitancy Scale

|  | EFA loadings<br>(n=502) |  | CFA standardized regression<br>weights (n=531) |  |
| --- | --- | --- | --- | --- |
|  | VHS factor 1:<br>Lack of<br>confidence | VHS<br>factor 2:<br>Risks | VHS factor 1:<br>Lack of<br>confidence | VHS<br>factor 2:<br>Risks |
| Vaccines are important for my health (VHS) | 0.902 | 0.166 | 0.87 |  |
| Vaccines are effective (VHS) | 0.877 | 0.118 | 0.87 |  |
| Being vaccinated is important for the health of<br>others in my community (VHS) | 0.907 | 0.139 | 0.85 |  |
| All vaccines offered by the government<br>programme in my community are beneficial (VHS) | 0.891 | 0.189 | 0.84 |  |
| New vaccines carry more risks than older vaccines<br>(VHS) (R) | -0.029 | 0.583 |  | 0.61 |
| The information I receive about vaccines from the<br>vaccine program is reliable and trustworthy (VHS) | 0.785 | 0.222 | 0.75 |  |
| Getting vaccines is a good way to protect myself<br>from disease (VHS) | 0.902 | 0.161 | 0.86 |  |
| Generally, I do what my doctor or health care<br>provider recommends about vaccines (VHS) | 0.822 | 0.18 | 0.76 |  |
| I am concerned about serious adverse effects of<br>vaccines (VHS) (R) | 0.141 | 0.789 |  | 0.74 |
| I am uncomfortable getting a vaccine that was<br>rushed into production (Seale et al., 2010) (R) | -0.018 | 0.727 |  | 0.48 |
| I feel uncomfortable getting vaccinated<br>(Maisonneuve et al., 2018) (R) | 0.344 | 0.724 |  | 0.72 |
| Government over hypes the need for vaccines<br>(Gidengil et al., 2019) (R) | 0.338 | 0.735 |  | 0.33 |
| Corporations manufacturing vaccines only care for<br>profit (Gidengil et al., 2019) (R) | 0.175 | 0.641 |  | 0.55 |
| Vaccines cause diseases (Gidengil et al., 2019) (R) | 0.407 | 0.689 |  | 0.74 |

*Note:* Items were coded on a 5-point scale, from *strongly agree* to *strongly disagree*—higher scores indicate higher vaccine hesitancy. EFC refers to exploratory factor analysis; CFA refers to confirmatory factor analysis; (R) refers to recoded items; VHS refers to Vaccine Hesitancy Scale (e.g., Luyten et al., 2019). For EFA, factors were extracted using Principal Component Analysis with varimax rotation (with Kaiser Normalization).

Confirmatory factor analysis assessed the vaccine hesitancy scale using items from the other half of the sample. The best goodness-of-fit for the 14-item version was with two factors (RMSEA = 0.078, CFI = 0.95, TLI = 0.94). Thus, a two-factor structure, consisting of a “lack of confidence” (*r’s* = 0.62 to 0.82, *α =* .95) part with 7 items, and a “risks perception” (*r’s* = 0.28 to 0.63, *α =* .82) part with 7 items, showed the best psychometric characteristics of the VHS. On a scale from 1 to 5, with a higher score indicating higher vaccine hesitancy, the average respondent scored 1.97 (SD = 0.98) for the lack of confidence factor and 2.89 (SD = 0.93) for risks, highlighting that the hesitancy was driven more by risk perceptions than by lack of confidence in vaccines.

### Intension to get a COVID-19 vaccine

Several demographic variables were correlated with willingness to get vaccinated against COVID-19 in bivariate analyses (See Table 3). Men compared to women, older respondents compared to younger, more educated compared to less educated, higher annual income compared to lower-income, Asian and other ethnicities compared to Māori, respondents without children compared to those with children, and those who did not smoke or vape compared to those who do, were more likely to say they intend to get vaccinated against COVID-19 when one becomes available.

**Table 3.** Bivariate Correlates of COVID-19 Vaccine Intention with Categorical Variables

|  | *N* = 1040 | Intention to<br>vaccinate against<br>COVID-19<br>(Yes %) | Bivariate RR<br>(95% CI) |
| --- | --- | --- | --- |
| *Sex* |  |  |  |
| Male | 510 | 78% | 1.10 (1.03 to 1.18)** |
| Female | 530 | 70% | Ref |
| *Age* |  |  |  |
| 18-25 | 146 | 72% | 0.89 (0.79 to 1.00)* |
| 26-35 | 187 | 70% | 0.88 (0.80 to 0.98)* |
| 36-45 | 166 | 71% | 0.89 (0.79 to 0.99)* |
| 46-55 | 187 | 72% | 0.90 (0.81 to 1.01) |
| 56-65 | 156 | 76% | 0.94 (0.84 to 1.05) |
| 66 and above | 198 | 81% | Ref |
| *Education* |  |  |  |
| No qualification | 199 | 71% | 0.88 (0.8 to 0.97)* |
| Level 1 to Level 6 diploma | 564 | 71% | 0.86 (0.8 to 0.93)*** |
| Bachelor's degree or higher | 277 | 81% | Ref |
| *Ethnicity* |  |  |  |
| European New Zealander | 640 | 76% | 0.97 (0.89 to 1.06) |
| Māori | 170 | 64% | 0.83 (0.72 to 0.94)** |
| Pasifika | 80 | 69% | 0.91 (0.77 to 1.08) |
| Asian or Other | 150 | 80% | Ref |
| *Annual personal income* |  |  |  |
| Up to \$39,999 | 559 | 70% | 0.90 (0.82 to 0.99)* |
| \$40,000 to \$79,999 | 318 | 78% | 1.01 (0.92 to 1.11) |
| \$80,000 and above | 160 | 78% | Ref |
| *Employment status* |  |  |  |
| Other | 530 | 73% | 0.95 (0.89 to 1.03) |
| Currently employed | 510 | 75% | Ref |
| *Parental status* |  |  |  |
| Other | 421 | 77% | 1.07 (1.01 to 1.15)* |
| Has children | 619 | 72% | Ref |
| *Smoking status* |  |  |  |
| No | 738 | 76% | 1.09 (1.00 to 1.18)* |
| Yes | 302 | 70% | Ref |

Note. RR = relative risk; CI = confidence interval; Ref = reference group. \* $p < 0.5$ , \*\* $p < .01$ , \*\*\*
$p < .001$ .

In multivariable analyses (See Table 4), men compared to women (RR = 1.11, 95% CI: 1.05–1.18) and those without children compared to those with children (RR = 1.09, 95% CI: 1.02–1.16) were more willing to get a COVID-19 vaccine. Participants were also more willing if they reported higher levels of trust in scientific experts (RR = 1.10, 95% CI: 1.04–1.16). Moreover, with respect to general vaccine hesitancy attitudes, respondents with higher confidence about vaccines (RR = 0.77, 95% CI: 0.73–0.83) and those who perceived fewer risks (RR = 0.91, 95% CI: 0.87–0.96) were more willing to get a COVID-19 vaccine. There was no significant association between COVID-19 knowledge and intention to get vaccinated against COVID-19.

**Table 4.** Multivariable Correlates of COVID-19 Vaccine Intention

|  | Multivariable RR (95% CI) |
| --- | --- |
| *Sex* |  |
| Male | 1.11 (1.05 to 1.18)** |
| Female | Ref |
| *Age* |  |
| 18-25 | 0.93 (0.83 to 1.05) |
| 26-35 | 0.98 (0.87 to 1.10) |
| 36-45 | 0.94 (0.83 to 1.05) |
| 46-55 | 0.97 (0.87 to 1.07) |
| 56-65 | 0.92 (0.83 to 1.02) |
| 66 and above | Ref |
| *Education* |  |
| No qualification | 1.02 (0.93 to 1.12) |
| Level 1 to Level 6 diploma | 0.96 (0.9 to 1.03) |
| Bachelor's degree or higher | Ref |
| *Ethnicity* |  |
| European New Zealander | 0.95 (0.87 to 1.04) |
| Māori | 0.95 (0.84 to 1.07) |
| Pasifika | 1.02 (0.87 to 1.2) |
| Asian or Another Category | Ref |
| *Annual personal income* |  |
| Up to \$39,999 | 1.01 (0.91 to 1.12) |
| \$40,000 to \$79,999 | 1.02 (0.93 to 1.12) |
| \$80,000 and above | Ref |
| *Employment status* |  |
| Other | 0.97 (0.9 to 1.04) |
| Currently employed | Ref |
| *Parental status* |  |
| No children | 1.09 (1.02 to 1.16)* |
| Has children | Ref |
| *Smoking status* |  |
| No | 0.95 (0.88 to 1.02) |
| Yes | Ref |
| Trust scientists | 1.09 (1.04 to 1.16)** |
| Knowledge about COVID-19 | 0.99 (0.97 to 1.03) |
| *Vaccine hesitancy* |  |
| Lack of confidence | 0.77 (0.73 to 0.83)*** |
| Risks | 0.91 (0.87 to 0.96)*** |

Note. RR = relative risk; CI = confidence interval; ref = reference group. \* $p < 0.5$ , \*\* $p < .01$ , \*\*\*
$p < .001$ .

## Discussion

A national sample survey in New Zealand shows that 74% of adults intend to get a COVID-19 vaccine when one becomes available. This is similar to findings from global surveys (O’Keefe, 2020), although slightly higher than several countries such as Australia (Rhodes et al., 2020), US (Reiter et al., 2020), and several European countries (Neumann-Böhme et al., 2020).

The most novel finding of this study is that a generalized measure of vaccine hesitancy is significantly associated with COVID-19 vaccine intention. The vaccine hesitancy scale with 14-items was developed following the 9-item VHS scale for the general population (Luyten et al., 2019), with additional items sourced from systemic reviews of vaccine hesitancy (Gidengil et al., 2019; Larson et al., 2015, 2018; Maisonneuve et al., 2018; Verger & Dubé, 2020). In probably the first study of its kind, vaccine hesitancy scale was tested in New Zealand, adding to few studies of vaccine hesitancy outside Europe and the US (Larson et al., 2018). For example, (Seale et al., 2010) found that the perception of rushed vaccine production was one of the most cited fears for the H1N1 vaccine intention. This measure is particularly important given that public perceptions of a rushed vaccine development or inadequate testing may lead them to question the safety of the vaccine and increase distrust with vaccination in general (Hoffman, 2020). Findings of this study can help standardize the VHS scale to advance research on vaccine hesitancy and its enduring impact on public attitudes towards new vaccines.

Knowledge about COVID-19 was not associated with COVID-19 vaccine intention. This finding is worrying as another study in the US also shows that knowledge about COVID-19 infection was only moderately associated with COVID-19 vaccine acceptance in bivariate analysis and was not significantly associated in the presence of other covariates (Reiter et al., 2020). Knowledge about a disease origin and prevalence alone may be an insufficient motivator for individual action, indicating a need for future studies to replicate these findings.

Trust in scientific experts is one of the strongest correlates of COVID-19 vaccine intention, similar to previous studies on general vaccination behaviors (e.g., de Figueiredo et al., 2020). Individuals rely on trusted sources of information to make judgments in uncertain information context, such as the rapid, multinational effort to develop a COVID-19 vaccine. Building public trust in scientific experts, particularly among historically disenfranchised communities who may already harbor high distrust with government and scientific elite, by building community relationships is particularly important to decrease the vaccination gap among the most vulnerable groups.

The strengths of this study include a large representative sample, multiple items to measure and test vaccine hesitancy, trust in scientific experts, and knowledge about COVID-19. A primary limitation is the cross-sectional design of the study. Attitudes towards COVID-19 vaccine are likely to shift, as seen in other countries (e.g., Rhodes et al., 2020), therefore a need to further monitor shifting public attitudes and correlates affecting this shift. Future research should also explore other dimensions of vaccine hesitancy, such as the desire for autonomy and morality concerns (Gidengil et al., 2019).

### Conclusion

The World Health Organization identifies vaccine hesitancy as one of the 10 most important health threats to the world (World Health Organization, 2019). Increasing public enthusiasm for vaccination should co-occur with the development of a COVID-19 vaccine. Findings of this study suggest that a health communication campaign from trusted sources, with information that addresses prevailing concerns about vaccines, is likely to help increase COVID-19 vaccine uptake.

## Data Availability

Data will be made available after peer-reviewed publication

